# Assessment of the school environment for risk factors for tungiasis in nine counties of Kenya: a cross-sectional survey

**DOI:** 10.1101/2024.11.02.24316660

**Authors:** Lynne Elson, Christopher Kamau, Sammy Koech, Christopher Muthama, George Gachomba, Erastus Sinoti, Elwyn Chondo, Eliud Mburu, Miriam Wakio, Jimmy Lore, Marta Maia, Ifedayo Adetifa, Benedict Orindi, Phillip Bejon, Ulrike Fillinger

## Abstract

**Background:** Tungiasis is a neglected tropical skin disease mostly affecting children under 15 years, the elderly and disabled people in the most resource poor populations in Latin America and Sub-Saharan Africa. While most transmission seems to occur at home inside houses, we aimed to identify factors in the school environment that may put children at increased risk of infection.

**Methods:** As part of a cross-sectional school-based prevalence survey of 21,466 pupils in nine counties of Kenya, observations of school infrastructure were made and the headteacher interviewed in each of 196 schools. In a subset of 97 schools, detailed observations of 322 classrooms were made and 117 teachers interviewed. Mixed effect logistic regression models were used to identify factors associated with tungiasis infection in pupils.

**Results:** We found a higher odds of tungiasis infection for pupils in schools where more than 400 pupils were enrolled (aOR 2.24, 95% CI 1.12−4.50, p=0.023), where clean water was not always available (aOR 2.28, 95% CI 1.13−4.60, p=0.021); and if the school buildings were in a bad structural condition (aOR 2.15, 95% CI 1.00−4.62, p=0.050). Of the 5,102 pupils in 97 schools, 99% studied in a classroom with a concrete floor and those with a lot of loose soil or sand on top of the concrete floor had a six times higher odds of infection than those on a clean concrete floor (aOR 6.52, 95% CI 1.61−26.35, p=0.008). Only 45% of (head)teachers in affected schools knew they had infected pupils in their school or grade. Those who did, were aware of the impact it was having on the pupils and yet only three of 76 affected schools had conducted a tungiasis intervention activity within the last year.

**Conclusion:** Most schools do not pose a risk of tungiasis for pupils, instead the home environment is the main risk. However, where school buildings are not well maintained and water not always available a multisectoral approach to control tungiasis is needed involving the Department of Health as well as the school management and the Department of Education.

## Background

Tungiasis is a skin disease mostly affecting children under 15 years, the elderly and disabled people in the most resource poor populations in Latin America and Sub-Saharan Africa[1]. It has been neglected by governments, donors, and the scientific community to this day, and was only recently added to the World Health Organization’s list of neglected tropical disease[2] and neglected diseases of the skin [3]. The disease is caused by the sand flea, *Tunga penetrans.* The female of this species burrows into the skin, where it remains embedded for the duration of its five-week adult life, unless removed by its host [4]. The flea is less than 1mm in length when it seeks out its host, once in the skin it copulates with the free-living male flea and grows 2000 times in size as eggs develop in the abdomen. Hundreds of eggs are released into the environment, where larvae will hatch and develop in the upper layer of soil and dust if conditions are suitable. After one week the larvae pupate, cloaking themselves in sand grains, and after one more week the adult emerges [5].

The rapid growth of the embedded female in the skin causes considerable pain and the flea induces an inflammatory response resulting in intense itching, often exacerbated by secondary bacterial infection [6, 7]. A few studies have demonstrated tungiasis reduces a patient’s quality of life through reduced mobility, disturbed sleep, concentration at school or work, stigma and discrimination [8, 9]. It was also shown that tungiasis was associated with higher absenteeism and lower school performance [8].

People’s own homes have been identified as the most likely source of infection through risk factor studies of households [10] and schools [11], and by identification of the larvae in the soils of unsealed earthen floors in the sleeping rooms of houses [5, 12]. However, since the most affected population are children under 15 years, it might be expected that schools, which has a high density of children for a large proportion of each day, may contribute to the risk of infection. The objective of this study was to determine whether any aspects of the school environment may be associated with tungiasis infection among pupils and explore headteachers’ and teachers’ knowledge and attitudes relating to risk factors for, and the impact of tungiasis on their pupils.

## Methods

### Study design and population

This study was a comparison of school-related risk factors for infected and uninfected pupils, nested in a cross-sectional survey in nine counties which was designed to determine the national prevalence of tungiasis in Kenya [13]. The counties were purposively selected to represent the five major climatic zones, cultures and geographies of the country: Turkana, Samburu, Kericho, Nakuru, Muranga, Kajiado, Makueni, Taita Taveta and Kilifi.

### Sample size and selection

The number of pupils and schools to enrol was based on the sample size of pupils needed for the national prevalence estimate. A total of 21,466 pupils were enrolled from 196 schools across the nine counties. Within each county 22 primary schools were stratified randomly selected from pre-existing school lists for each sub-county, selecting at least two schools from each sub-county. The schools were surveyed between May 2021 and April 2023. Within each selected school 114 pupils aged 8 to 14 years, equal number of girls and boys, were examined for tungiasis [13].

### Procedures

Observations were conducted of the school infrastructure including the composition and condition of buildings, washing and sanitation facilities, and presence of animals on the compound. For half of the schools, observations were also made of individual classrooms: measuring the length and width of the classroom, obtaining enrolment and absentee numbers for the day of the visit, the materials used for construction, the condition of the classroom floor and building and the amount of loose soil on a concrete floor. In every school, the headteacher or their representative was interviewed, and in half of the schools up to eight teachers per school, using a semi-structured questionnaire regarding themselves, the school and pupil welfare. Headteachers and teachers who said they currently or previously had tungiasis cases in the school were asked questions about their knowledge and attitudes of tungiasis and infected pupils. Where questions were open, limited space was provided for responses. The tool was piloted, and necessary adjustments made before implementation. The questionnaire was pre-translated and administered in Kiswahili. The field interviewers were trained by the lead investigator. All interview responses were entered into electronic tablets using the REDCap web application [14] and uploaded to the Kenya medical Research Institute (KEMRI)-Wellcome Trust servers.

### Observations

The outcome of interest was a pupil’s infection status defined as tungiasis case if they had at least one embedded flea (live, dead or manipulated) on their hands or feet, and uninfected otherwise.

The selection of explanatory variables for the regression models was based on the biology of the parasite. For instance, buildings in poor condition, unsealed earthen floors, concrete floors that are broken/ cracked and/ or covered with loose soil or dust and general poor cleanliness might be expected to support the development of off-host stages of *T. penetrans*. Previously it has been shown that adult female fleas can jump from one heavily infected host to another in proximity [15], so we included the total pupil enrolment in a school and the density of pupils in a classroom. Past studies have identified factors relating to hygiene and sanitation to be associated with disease, therefore access to water, washing stations, presence of hygiene promotion signs, access to toilets and the condition of toilets were included in the observations. The attitudes of headteachers and teachers towards pupil welfare was explored as it could potentially impact the vulnerability of pupils for infection. In this context, questions were asked around supervision of pupil hygiene behaviors and shoe-wearing, the actions taken if pupils do not match the school standards, and whether regular hygiene promotion, deworming or jigger programs were conducted. Since county, age and sex have previously been shown to be important factors associated with tungiasis in this study, these were also included in the models.

### Data Analysis

#### School and classroom-related risk factors

Mixed effects logistic regression models were used to identify factors related to the school and classrooms that may be associated with pupils’ tungiasis infection status (pupil infected or not). Fixed effects included the county, pupil age and sex, school and classroom characteristics described in the observations paragraph above. The school was used as a random effect to control for the clustered study design. All variables with a p-value less than 0.2 in univariable models were included in a multivariable model. Backward elimination, sequentially excluding variables was used for the final model. Akaike information criteria (AIC) was used to compare the models. Models with lower AIC values were preferred. Additional consideration on variable selection was expert knowledge on potential relationships with the outcome. Results are presented as odds ratios or adjusted odds ratios with 95% confidence intervals and p-values.

#### Headteacher and teacher interviews

Pearson Chi-square was used to test whether headteachers’ awareness of the tungiasis status of pupils (said yes to question “are there any tungiasis cases in your school”) was different to the actual status established during foot examinations. The same was conducted for teachers’ awareness of tungiasis cases in the grade that they taught most of the time. Wilcoxon rank-sum tests were used to test whether their awareness was related to the prevalence of tungiasis in their school. The remaining responses of the interviews were summarized using the percent of respondents with each response type.

All analyses were conducted using Stata BE version 18 (Stata Corp LLC, College Station, Texas, USA). All tests were performed at 5% significance level.

## Results

### Study population

A total of 196 schools were enrolled in the study and 21,466 pupils were examined, of whom 242 (1.1%) were found to be infected. The infected pupils were from 76 schools (“affected schools” in Fig 1). In all 196 schools, observations were made of the school infrastructure and enrollment records and headteachers or their representatives were interviewed. For a subset of 97 schools, 35 of whom had tungiasis cases, detailed observations were made of 322 classrooms and 117 teachers were interviewed (Fig 1). Within the 76 affected schools, the school prevalence of tungiasis varied from a low of 0.8% to a high of 22.3% among pupils aged 8 to 14 years. The distribution of pupils by all explanatory variables is presented in the Additional file S1.

**Figure 1.**
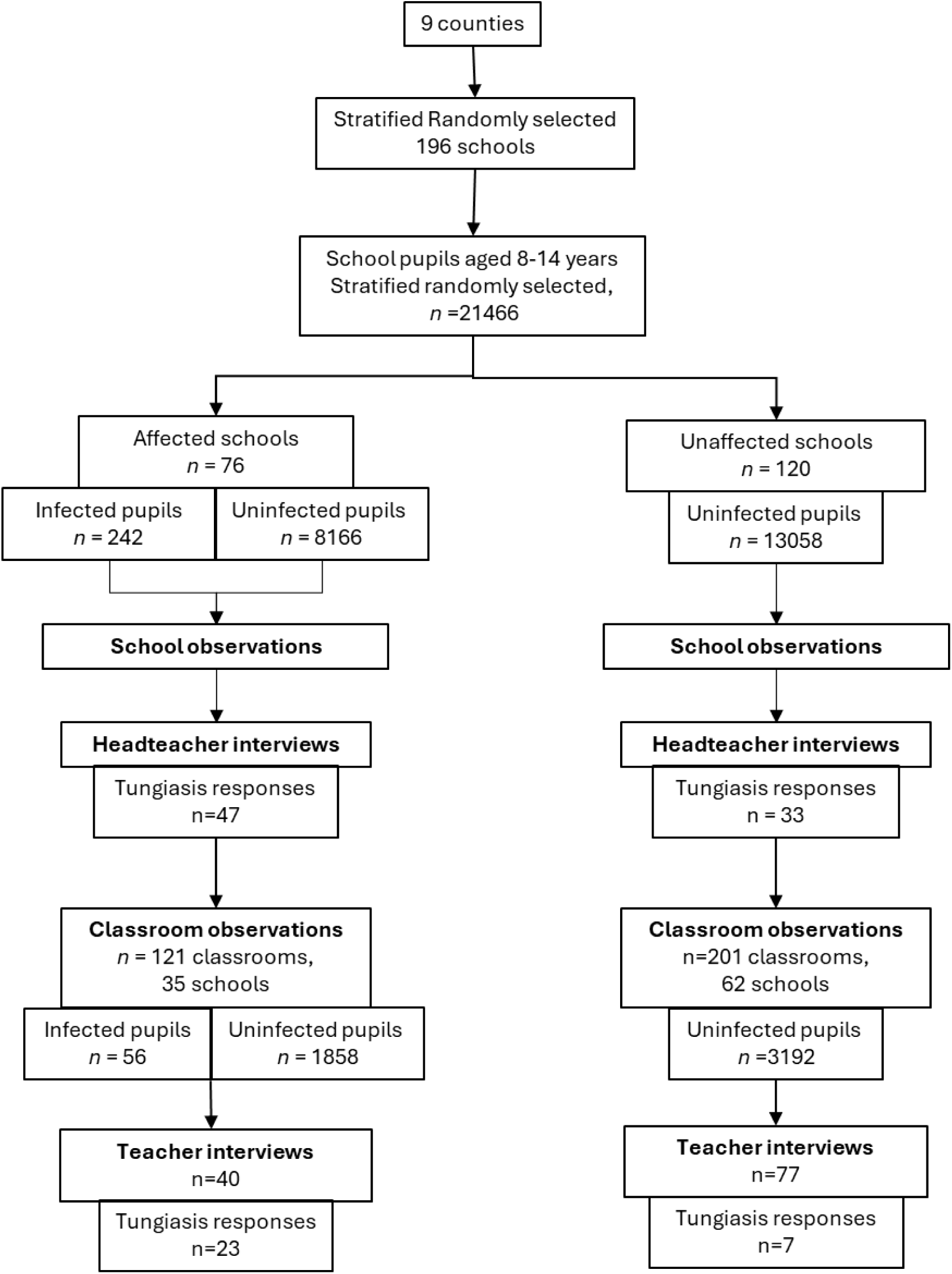
Flow chart of participants and activities. A school was considered “affected” if at least one case was identified during examinations of 114 randomly selected pupils aged 8 to 14 years. n: number. “Tungiasis responses” refers to the number of headteachers or teachers who responded to questions regarding tungiasis in the interview.

### School-related risk factors

During univariable analysis, many of the observed factors were associated with tungiasis (Table 1) and were included in the multivariable analysis. In the final model, pupil age and sex were associated with tungiasis status and only a few school factors. Pupils were twice as likely to be infected in a school where more than 400 pupils were enrolled (aOR 2.24, 95% CI 1.12−4.50, p=0.023); where clean water was not always available (aOR 2.28, 95% CI 1.13−4.60, p=0.021); that had a mass deworming event within one to three months prior to the tungiasis survey (aOR 2.06, 95% CI 1.03−4.16, p0.042); and although borderline, if the school buildings were in a bad structural condition (aOR 2.15, 95% CI 1.00−4.62, p=0.050). While not significant, the ratio of boys to girls in the school was retained in the model based on the AIC and Wald tests indicating pupils in schools with a higher ratio of boys to girls had four times the risk of infection (aOR 4.27, 95% CI 0.78−23.27, p=0.093).

**Table 1.**
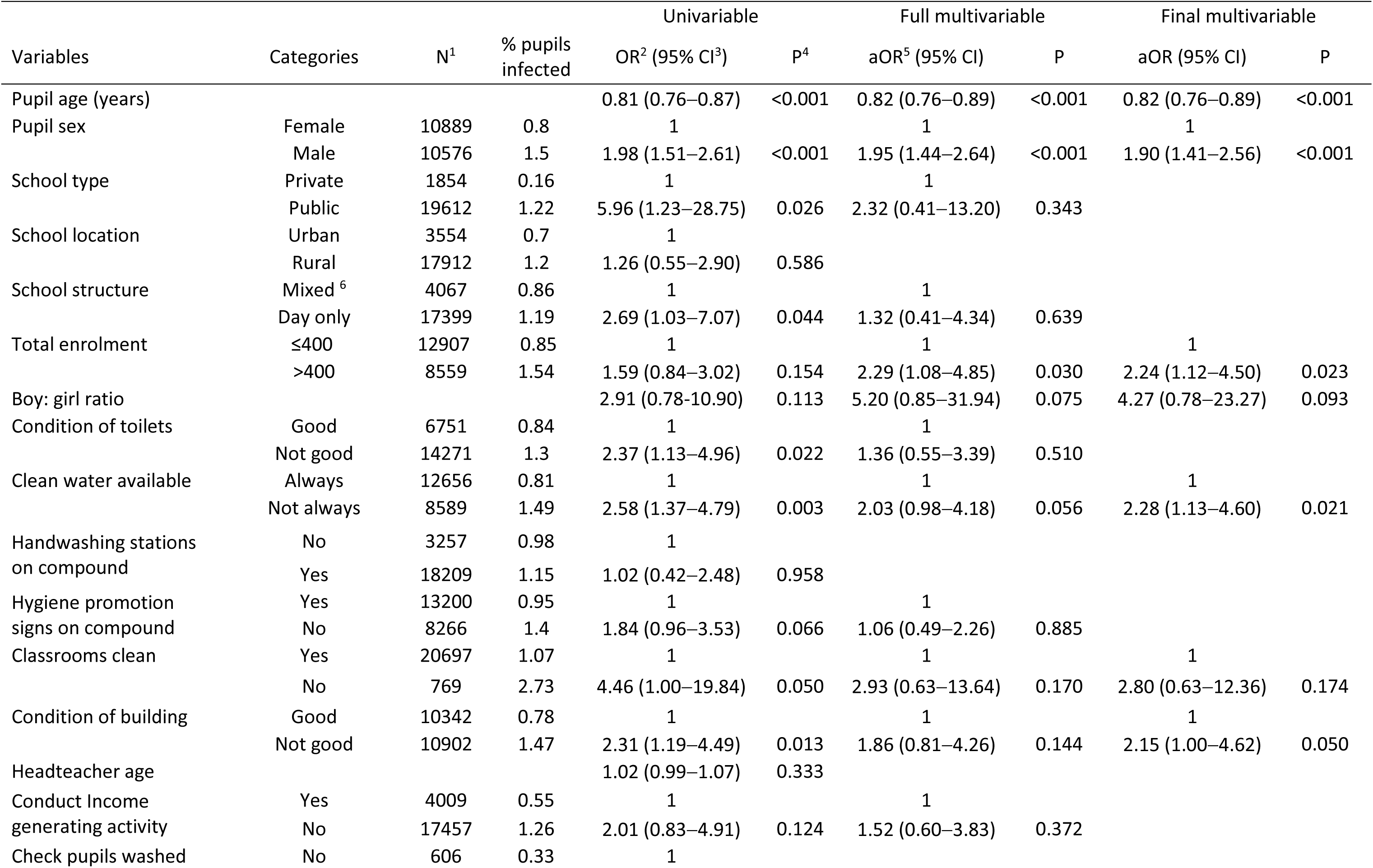
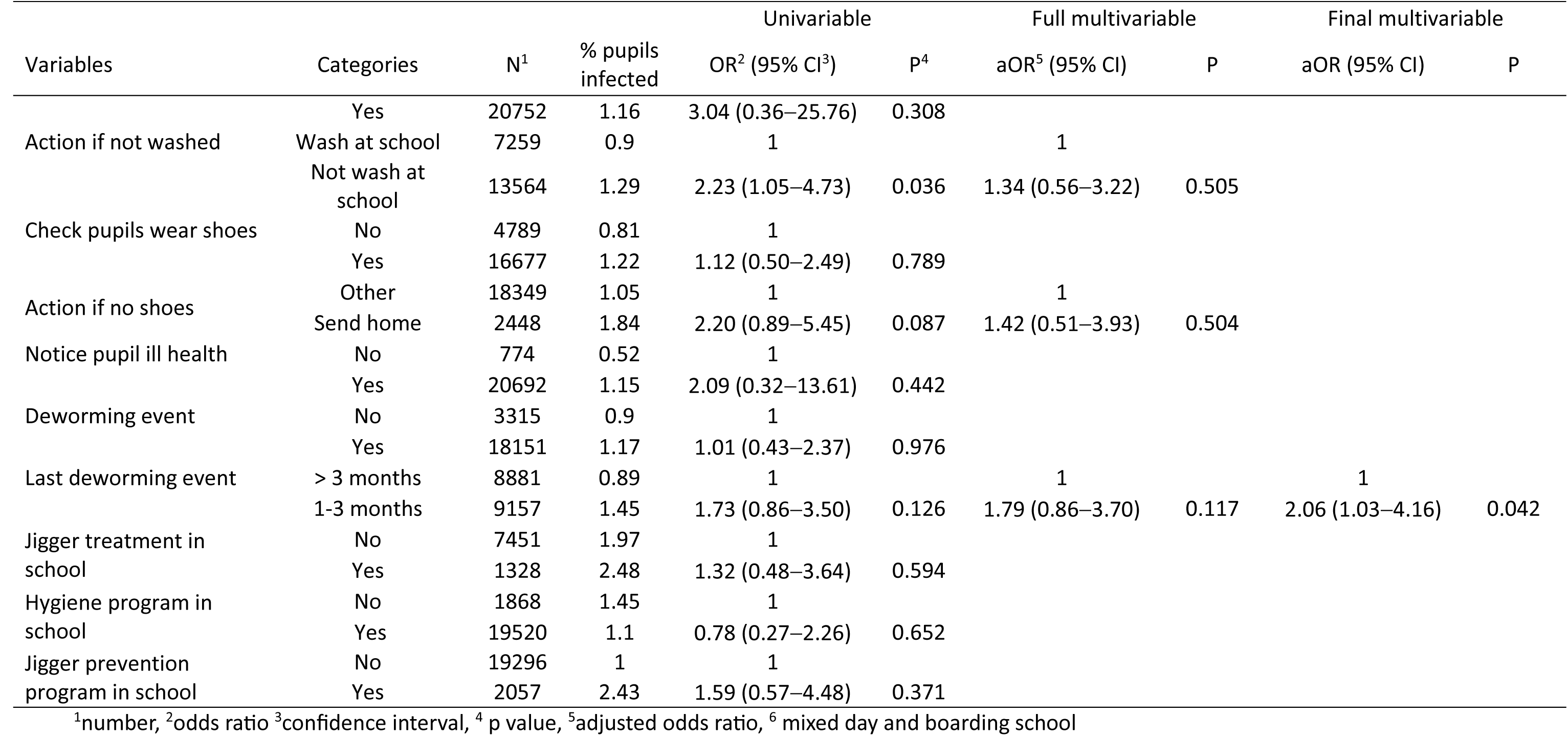
Mixed effects logistic regression of school-related factor for tungiasis infection in pupils, county and school as random effect (n=21,465, 242 cases).

### Classroom-related factors

Specific classroom factors were collected in 97 schools for 322 classrooms of which 40 (12.4%) classrooms had at least one case. A total of 5,106 pupils studied in the 322 classrooms, of whom 56 (1.1%) were infected. Although the construction materials of the classrooms were recorded, these were not included in the models since 99.4% of classrooms had an iron sheet roof, 95.6% had walls of bricks, cement blocks or stones and 99% had concrete floors. The distribution of pupils by all classroom variables is presented in the Additional file S2.

In the univariable analysis, tungiasis was positively associated with all the variables assessed except the number of pupils enrolled in a classroom and the pupil density (number of pupils per m^2^) in a classroom (Table 2). The condition of the concrete floor and cleanliness of the classroom were not included in the final multivariable model since they were conceptually related and positively correlated to the amount of loose soil or sand on the floor (Additional file S3, Pearson’s chi2 p<0.001). Pupils who studied in a class with a lot of loose soil or sand on the floor had a six times higher odds of being infected (aOR 6.52, 95% CI 1.61−26.35, p=0.008) albeit with a wide confidence interval due to the low number of cases (Table 2). Tungiasis was also associated with the number of pupils absent on the day of the survey (aOR 1.11, 95% CI 1.01−1.22, p=0.023). As before, pupil sex was also associated with infection.

**Table 2.**
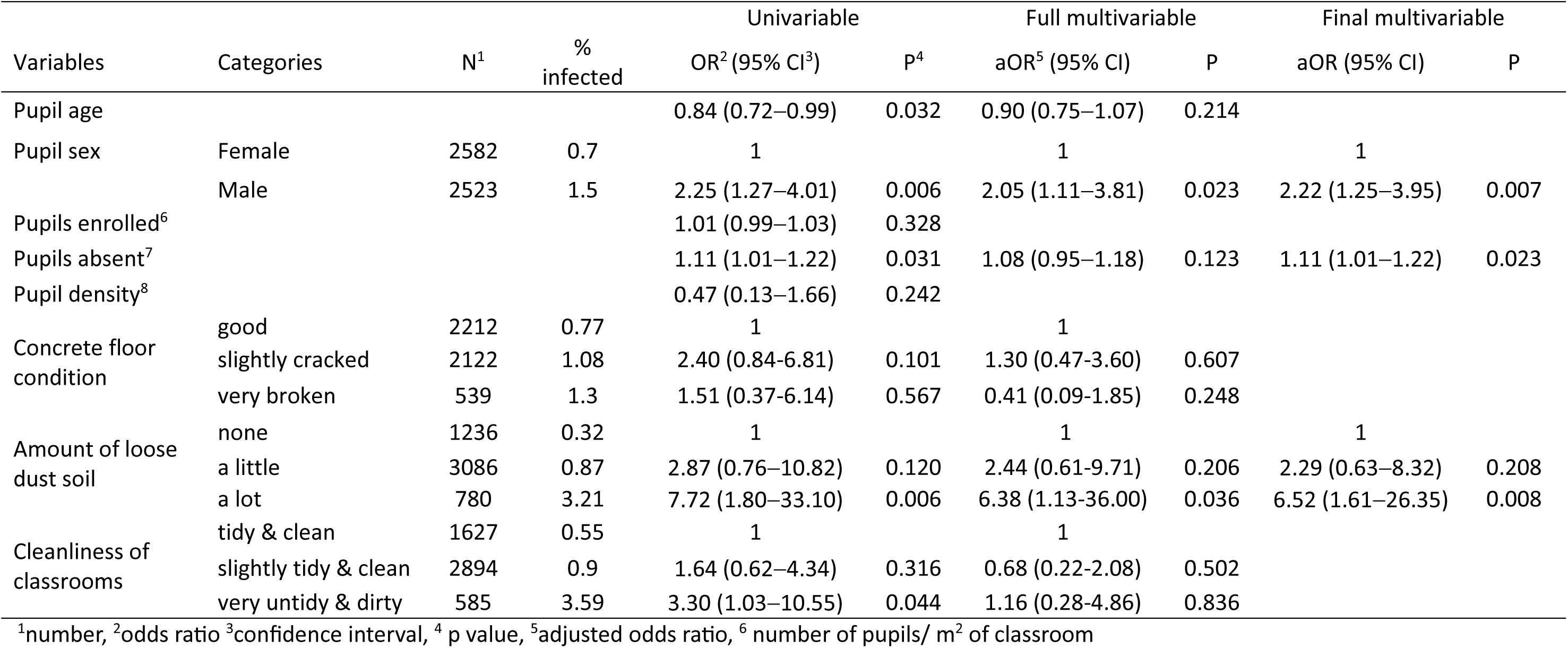
Mixed effects logistic regression for classroom factors associated with pupil infection, county and school as random effects (n=5102 pupils, 56 infected).

### Headteachers and teachers’ knowledge and attitudes

Of the 197 headteachers interviewed, 26.4% (52) were female, while 51% (60) of the 117 teachers were female. Both teachers and headteachers had worked in the school for a median of three years (IQR 2-6).

#### Head (Teacher) awareness of tungiasis situation in schools

To assess the level of awareness of the tungiasis status of pupils in their school, headteachers were asked if they thought there were cases in the school, and we compared their response to the results of our foot examinations. At the time of the interview respondent headteachers were not aware of the results of the pupil examinations conducted by the research team. Of the 76 headteachers in schools with at least one case, only 45% (34) were correct in reporting there were cases in their school (Pearson Chi^2^ p <0.001, Table 3). Teachers were asked if they knew of cases in their grade. Of the 19 teachers in grades with at least one case, 74% (14) were correct in reporting they had cases in their grade (Table 4).

**Table 3.** Headteacher awareness of the tungiasis situation in their school.

| Headteachers | Actual foot examination results | | | Pearson $\chi^2$ p value |
| --- | --- | --- | --- | --- |
|  | At least one case in the school | No cases in the school | Total |  |
| Yes, have cases in my school | 34 (44.7%) | 9 (7.5%) | 43 (21.9%) | <0.001 |
| No cases in my school | 42 (55.3%) | 111 (92.5%) | 153 (78.1%) |  |
| Total | 76 | 120 | 196 |  |

**Table 4.**
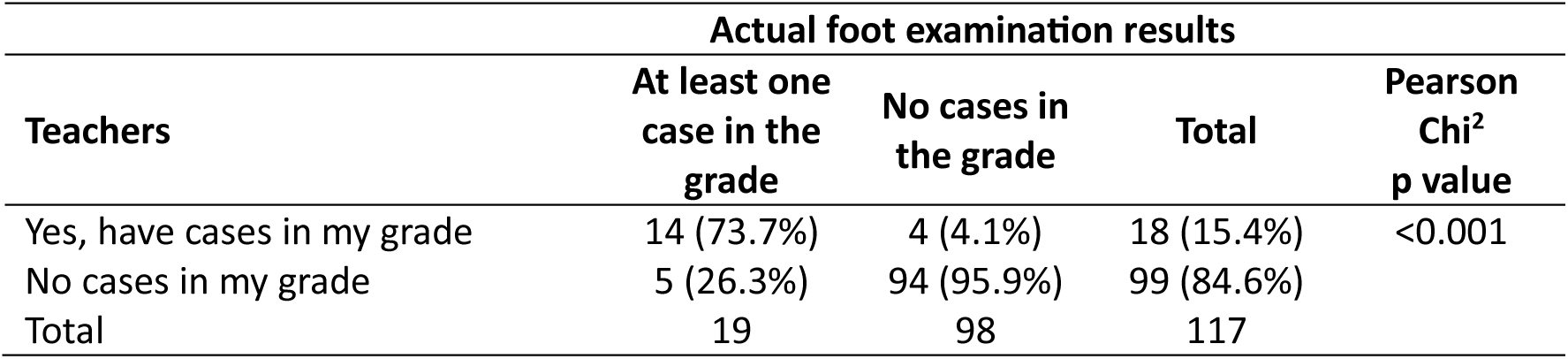
Teacher awareness of the tungiasis situation in their grade.

| Teachers | Actual foot examination results |  |  | Pearson<br>Chi<sup>2</sup><br>p value |
| --- | --- | --- | --- | --- |
|  | At least one<br>case in the<br>grade | No cases in<br>the grade | Total |  |
| Yes, have cases in my grade | 14 (73.7%) | 4 (4.1%) | 18 (15.4%) | <0.001 |
| No cases in my grade | 5 (26.3%) | 94 (95.9%) | 99 (84.6%) |  |
| Total | 19 | 98 | 117 |  |

The 55% of headteachers who incorrectly reported that the school had no cases, were in schools with a lower prevalence (median 0.95%, IQR 0.9−2.7) than those who were correct (median 2.25%, IQR 0.9−4.4, p=0.065). However, this difference was not statistically significant in the Wilcoxon rank sum test. For teachers the difference was significant (median 2.7%, IQR 1.8−5.4 vs. 0.9%, IQR 0.9−1.8, p<0.001).

One limitation often proposed for using school-based surveys to assess prevalence and risk factors, is that the most severe cases will be missed as they are unable to attend school on account of their poor mobility. To date there is no evidence to support this, so we asked teachers if they knew if there were pupils missing from their class due to tungiasis, and four teachers from four schools (11.4% of affected schools) said there were. The schools with pupils absent on account of tungiasis had higher prevalence among the pupils who did attend, than schools who did not have any pupils absent on account of tungiasis (median 4.6, IQR 2.3−7.1 vs. 1.8, IQR 0.9−3.1, Wilcoxon rank sum p=0.002).

#### Knowledge of and impact of tungiasis

Those headteachers and teachers who said they currently or previously had tungiasis cases in their school were asked more questions about the disease and its impact on pupils. A total of 80 headteachers and 30 teachers responded (Fig 1). The headteachers and teachers were asked why they thought infected pupils had tungiasis and the majority attributed it to family poverty (70%) and poor hygiene (87%). A few also suggested it was due to lack of adult care (15.5%), lazy parents (18.2%), and to living in an area with red soil (14.5%), and 80% consider the disease to be seasonal, being worse during “hot, dry, and dusty” times of the year.

Respondents were asked their opinion as to how they thought pupils with tungiasis are affected by the disease. The most common impacts mentioned were poor concentration in class, low academic performance, poor attendance, mobility, stigma, “poor health/ sickly”, low self-esteem, and being “uncomfortable or restless”. Other impacts mentioned by a few respondents were being “tired”, sleeping in class, “mentally disturbed”, “poor memory”, and “socially isolated”.

When headteachers and teachers were specifically asked about the psychological impact on pupils compared to uninfected pupils, only two felt they were no different to uninfected pupils. Around one third mentioned that they were sad (29.1%), quiet (25.5%), withdrawn (36.4%) and seemed ashamed or embarrassed (38.2%). More than half (60%) said pupils with tungiasis are excluded by other pupils and 42.7% said pupils with tungiasis were ridiculed by others. When asked about the parents of the infected pupils, 58% felt the parents of infected pupils engage less, and 60% felt parents were less likely to come to school when asked, compared to parents of uninfected pupils.

### Health programs in school

Despite 21.9% (43) of headteachers thinking they had pupils in their school infected with tungiasis and being aware of the impact it had on their academic achievement, only 6.1% (12) of all headteachers reported they had had an intervention activity to treat the pupils, and for 75% (9) of those, that was more than a year before the interview. For the 12 schools who had had an intervention, only six had treated the pupils, either with potassium permanganate, coconut oil or neem oil and three had conducted, or provided chemicals for the fumigation of homes. Only 9.6% (19) of all headteachers reported having an active program to teach pupils about tungiasis and prevention, while 91.3% (179) of headteachers reported having a hygiene program and 60.7% (119) had other health interventions (COVID-19, polio, measles, trachoma, nutrition, menstrual hygiene, human papilloma virus and cervical cancer) in their school.

## Discussion

Given that those most affected by tungiasis are school aged children, it is often assumed that the school environment is a major risk factor for the spread of the disease. Apart from one other study conducted previously in Kilifi County in 2014 [11], no other study has investigated factors in schools that may be associated with tungiasis. Many risk factor studies have been conducted with school pupils in schools, but these have focused on the pupil and their home. In our own work, we previously concluded that the home environment is largely responsible for infection risk, but we also found indications that the school environment is a contributing factor [11]. Here we have investigated many more behavioral and environmental factors in depth but only a few factors were found to be associated with a pupil’s tungiasis infection.

We provide more evidence of an association of infection in pupils in schools with neglected infrastructure and poor cleanliness, having broken concrete floors and large amounts of loose soil or sand on the classroom floor. These factors are all linked to the developmental biology of the sand flea. The larvae need a certain amount of soil or sand to hide to prevent dehydration and avoid predation. The pupae require particles of soil/sand to coat themselves for protection [5]. Sand flea eggs expelled from the feet of an infected pupil wearing open or no shoes during class time, could drop on the sand and the off-host life cycle could be completed in the classroom. Consequently, keeping classroom floors in good condition and dust-free may help to reduce parasite development and pupil infection.

The positive association with having had a mass drug administration (MDA) event within the last three months for soil transmitted helminths (STH) and schistosomiasis is perhaps because the areas of the country targeted for frequent MDA, are also the areas in the country with the higher prevalence of tungiasis. Although integrated surveys have not yet been published, it might be expected that STH and tungiasis would have the highest prevalence in the same locations since they both tend to be found in children under the age of 15 years in resource-poor communities and the parasites all have an obligate life cycle stage in the soil from where infection is obtained [16]. The association also suggests the products and doses used in MDA are not effective against tungiasis.

Tungiasis has previously and in the current study population, been associated with not washing feet at least once a day with soap [10, 11, 13, 17]. Children in resource poor families may well not have access to water and/or soap at home, or parents lack the ability to enforce washing every day before and/or after school. We have found that in addition, if a school did not have constant access to water, the pupils had a higher risk of infection. Promoting foot hygiene and providing facilities for foot washing in school for those who do not have access at home could help reduce infection among pupils.

The majority of headteachers and teachers in affected schools knew that tungiasis was associated with poverty and poor hygiene and its impact on children, but there was rarely systematic action to help individual children to wash in school, nor implement a school-wide intervention program. While most schools had a general hygiene program, these programs focus on handwashing to prevent diarrheal diseases and soil transmitted helminths, and of course at the time of this study, COVID-19.

The association with a higher boy: girl ratio is not unexpected since boys in this study and previously have been shown [10, 18, 19] to be two to three times as likely to be infected than girls. Consequently, the more boys there are in a school the higher the risk for all pupils.

Often public health officials have relied on phoning school headteachers to determine which schools have pupils with tungiasis (LE personal experience). The findings of this study indicate headteachers are not very aware of the tungiasis situation in their school and are only likely to be if the school prevalence is over 1%. A higher proportion of teachers than headmasters correctly reported there were cases in the school or grade. This is not surprising since teachers have more contact with pupils each day, but there seems to be a lack of communication with school management to potentially address the problem.

Most headteachers and teachers felt tungiasis is seasonal and associated with the hot, dry or dusty times of year. Only one past survey in Brazil [20], has systematically investigated the seasonality and found the prevalence to be highest in the dry season. More studies are clearly needed to confirm this to provide health departments with the best time of the year to target for prevention interventions, before transmission begins to rise.

Most headteachers and teachers were not aware of the major impacts tungiasis has on children, except that they are stigmatized by other pupils, being excluded and ridiculed. Only a third or less of respondents in affected schools identified lack of concentration in class, absenteeism, poor academic performance, and the child’s psychological state. We have reported previously how the infected pupils in these schools, themselves reported having difficulty concentrating in class and sleeping at night, being ashamed, sad and angry, and observations of their school records found infected pupils missed more days of school and had lower exam results [8]. Many of these impacts have also been documented by previous studies [9, 21].

Something that has rarely been explored before in relation to children with tungiasis is the role parents may play in the child’s risk for infection. Two thirds of headteachers and teachers felt that parents of children with tungiasis were less engaged with the school than parents of uninfected pupils and were less likely to attend a meeting when specifically asked to come. This could reflect parental neglect of the child or shame regarding their child’s infection. Some teachers thought the child’s infection was caused by “lack of adult care”, or from having “lazy parents” and some were aware that there were problems at home for infected children and named poverty, alcohol use, divorce and illiteracy. The causes of neglect are likely to be many, but previously tungiasis has been shown to be associated with caregiver depression [22], which itself has been associated with child neglect [23] and poor hygiene of children [24]. Further research is needed to determine if there is an association between tungiasis and parental neglect, and to explore the possible reasons for child neglect in these communities and how that might be changed.

### LIMITATIONs

This study had one major limitation which was that it was not designed as a case: control study for risk factors, but rather was nested in a fully randomized prevalence survey which resulted in a low number of tungiasis cases encountered during the surveys, particularly when the detailed classroom observations and teacher interviews were conducted.

## Conclusions

Despite investigating many plausible environment and behavioral school-related factors, our study only found a few associated with tungiasis in pupils, reinforcing our previous conclusions that most risk is from pupils’ homes. In the few schools that have more than 400 pupils, classrooms in poor condition, cracked concrete floors not kept clean and tidy and free of loose soil or dust, do not have a constant supply of water, can contribute to the risk of tungiasis infection for their pupils. These findings reinforce the need for a one health, multisectoral approach to control tungiasis in a community, combining treatment with prevention which requires not only the Department of Health and health workers to target people in affected communities, but also the Board of Management of schools, teachers and the Department of Education to ensure all school buildings are in good condition, kept clean, and pupils encouraged to wash at school if they need to.

## Ethics approval and consent to participate

The study was approved by the KEMRI Scientific and Ethics Review Committee (approval number KEMRI/SERU/CGMR-C/170/3895) as well as the Oxford Tropical Research Ethics Committee (reference number 38-19). During the community engagement phase, a presentation was made to the national Director for Neglected Tropical Diseases at the Ministry of Health, the county and sub-County Health Management Teams and the Department of Education in all counties to obtain their approval. In each school a meeting was held with the school Parent Teachers’ Association (PTA) or management board to obtain their permission to conduct the survey in their school. The head teacher and PTA chairperson signed the consent form on behalf of the parents and school for the pupils to be examined. Each child gave verbal assent. Community health workers were hired and trained in each school to assist and be the link with the community emphasizing that participation was completely voluntary, and subjects had the opportunity to withdraw from the study at any point in the study.

For pupils selected for interviews, these were explained, and each was given an information leaflet to take home for their parents along with an opt-out form. Parents were to sign and return the form only if they did not want their child to participate in the interviews, or they could attend the school the next day to clarify any issues they may have. On the following day, if the selected pupils did not have the opt out form and were willing to participate, they proceeded with the interviews.

All data were collected on PIN protected electronic tablets, stored on password protected RedCap databases on the KEMRI-Wellcome Trust servers. Data were analyzed after export to Excel spreadsheets without inclusion of personal identifiers.

All pupils with tungiasis were referred for treatment to the community health workers or the local health facility using benzyl benzoate provided by the study. For those with secondary bacterial infection and other illnesses requiring treatment, a referral was made to the nearest health facility.

## Consent for publication

All the authors have reviewed and approved the publication of this paper.

## Availability of data and materials

The datasets and other materials supporting the conclusions of this article are available on KWTRP Research Data Repository at Harvard Dataverse through the following link: https://doi.org/10.7910/DVN/XM4K5I.

## Supplementary material

S1 Distribution of pupil population by school variables

S2 Distribution of pupil population by classroom variables

S3 Correlation of classroom variables with each other

## Competing interests

All authors declare no conflict of interest.

## Funding Information

Research funding for this work was provided by the Wellcome Trust through the project “Epidemiology of Tungiasis” (grant number 213724/Z/18/) granted to Lynne Elson as a Career Re-Entry Fellowship. This work was written with the permission of Director KEMRI-CGMRC. The views expressed herein do not necessarily reflect the official opinion of the donors. The funders had neither a role in the design of the study, nor in collection, analysis, interpretation of data nor in writing the manuscript.

## Author Contributions

Conceptualization, LE, UF, MM, IA, PB; Methodology, LE, UF, PB, BO, MM, IA; Formal Analysis, LE, BO; Investigation, LE, CK, SK, CM, GG, EC, MO, ES, JK, EM, MW, JL; Resources, LE, PB; Data Curation, LE ; Writing – Original Draft Preparation, LE.; Writing – Review & Editing, CK, SK, CM, GG, EC, MO, ES, JK, EM, MW, JL, UF, BO, PB, MM, IA; Visualization, LE.; Supervision, PB; Project Administration, LE, MM, PB; Funding Acquisition, LE, MM, PB.

## Data Availability

The datasets and other materials supporting the conclusions of this article are available on KWTRP Research Data Repository at Harvard Dataverse.

https://doi.org/10.7910/DVN/XM4K5I.

## Acknowledgements

We are grateful to the County Tungiasis Team Leaders: Wambani Zablon^3^, Collins Kipkorir^4^, Philip Nzyoka^5^, Moses Obiero^6^, Marawan Elijah^7^, Jacob Kapombe^8^, Kuyan Malano^9^, Caleb Kasuku^10^, Rael Kukule^11^ from Departments of Health in ^3^Muranga, ^4^Kericho, ^5^Makueni, ^6^Nakuru, ^7^Samburu, ^8^Kilifi, ^9^Kajiado, ^10^Taita Taveta, ^11^Turkana,

We are also grateful to the communities who participated, the school Parent Teacher Associations and Head Teachers who allowed us to work in their schools, the county Directors of Health and Education who gave their approval for the study and assisted in many ways to ensure its success. We acknowledge the efforts and dedication of Henry Kivuva (RIP) to set up the survey in Makueni County.

## Abbreviations

a*OR*: Adjusted odds ratio
CHMT: County Health Management Team
*CI*: Confidence Interval
KEMRI: Kenya Medical Research Institute
NTDs: Neglected Tropical Diseases
*OR*: Odds ratio
PTA: Parent Teachers Association
SES: Socio-economic status
*SD*: Standard deviation
WASH: Water, sanitation and hygiene

## Notes

### Competing Interest Statement

The authors have declared no competing interest.

### Author Declarations

The Scientific and Ethics Review Committee of KEMRI (approval number KEMRI/SERU/CGMR-C/170/3895) and the Oxford Tropical Research Ethics Committee (reference number 38-19) gave ethical approval for this work.

