## Supporting information for "Assessment of the school environment for risk factors for tungiasis in nine counties of Kenya: a cross-sectional survey"

### Elson Tungiasis supporting materials

#### S1 Distribution of pupil population by school variables

| Variables | Categories | Number missing | Uninfected Pupils N | Infected Pupils N (%) | Total | P value |
|---|---|---|---|---|---|---|
| **All** | | | 21224 | 242 (1.1) | 21466 | |
| County | Turkana | 0 | 2390 | 11 (0.5) | 2401 | <0.001 |
| | Samburu | | 2318 | 31 (1.3) | 2349 | |
| | Kericho | | 2396 | 14 (0.6) | 2410 | |
| | Muranga | | 2410 | 50 (2.0) | 2460 | |
| | Nakuru | | 2400 | 40 (1.6) | 2440 | |
| | Kajiado | | 2325 | 43 (1.8) | 2368 | |
| | Makueni | | 2426 | 13 (0.5) | 2439 | |
| | Taita taveta | | 2409 | 3 (0.1) | 2412 | |
| | Kilifi | | 2150 | 37 (1.7) | 2187 | |
| Pupil sex | Female | | 10804 | 85 (0.8) | 10889 | <0.001 |
| | Male | | 10419 | 157 (1.5) | 10576 | |
| School type | Day only | 0 | 17192 | 207 (1.2) | 17399 | 0.073 |
| | Mixed | | 4032 | 35 (0.9) | 4067 | |
| School support | Private | 0 | 1851 | 3 (0.2) | 1854 | <0.001 |
| | Public | | 19373 | 239 (1.2) | 19612 | |
| School location | Urban | 0 | 3528 | 26 (0.7) | 3554 | 0.014 |
| | Rural | | 17696 | 216 (1.2) | 17912 | |
| Headteacher sex | Male | | 15718 | 175 (1.1) | 15893 | 0.538 |
| | female | | 5506 | 67 (1.2) | 5573 | |
| Total number of pupils enrolled | ≤ 400 | | 12797 | 110 (0.9) | 12907 | <0.001 |
| | >400 | | 8427 | 132 (1.5) | 8559 | |
| Condition of buildings | Good | 222 | 10261 | 81 (0.8) | 10342 | <0.001 |
| | Not good | | 10742 | 160 (1.5) | 10902 | |
| Classrooms clean | No | 0 | 748 | 21 (2.7) | 769 | <0.001 |
| | Yes | | 20476 | 221 (1.1) | 20697 | |
| Condition of toilets | Good | 444 | 14086 | 185 (1.3) | 14271 | 0.004 |
| | Not good | | 6694 | 57 (0.8) | 6751 | |
| Access to clean water | Always | 221 | 12553 | 103 (0.8) | 12656 | <0.001 |
| | Not always | | 8461 | 128 (1.5) | 8589 | |
| Handwashing stations in school yard | No | 0 | 3225 | 32 (1.0) | 3257 | 0.395 |
| | Yes | | 17999 | 210 (1.2) | 18209 | |
| Hygiene signs in school yard | No | 0 | 8150 | 116 (1.4) | 8266 | 0.002 |
| | Yes | | 13074 | 126 (1.0) | 13200 | |
| Cats seen on compound | No | 110 | 19484 | 234 (1.2) | 19718 | 0.001 |
| | Yes | | 1632 | 5 (0.3) | 1637 | |
| Cows seen on compound | No | 110 | 15885 | 169 (1.1) | 16054 | 0.108 |
| | Yes | | 5231 | 70 (1.3) | 5301 | |
| Wild animals seen on compound | No | 221 | 19707 | 212 (1.1) | 19919 | 0.001 |
| | Yes | | 1299 | 27 (2.0) | 1326 | |
| Income generating project | No | 0 | 17237 | 220 (1.3) | 17457 | <0.001 |

| | | | | | | |
|---|---|---|---|---|---|---|
| | Yes | | 3987 | 22 (0.6) | 4009 | |
| Check pupils have washed | No | 108 | 604 | 2 (0.1) | 606 | 0.058 |
| | Yes | | 20512 | 240 (1.2) | 20752 | |
| Action if not washed | Assist at school | 643 | 7194 | 65 (0.9) | 7259 | 0.011 |
| | Not assist at school | | 13389 | 175 (1.3) | 13564 | |
| Check pupils wearing shoes | No | 0 | 4750 | 39 (0.8) | 4789 | 0.020 |
| | Yes | | 16474 | 203 (1.2) | 16677 | |
| Action if not wearing shoes | Other | 669 | 18157 | 192 (1.1) | 18349 | 0.001 |
| | Send home | | 2403 | 45 (1.8) | 2448 | |
| Notice pupil ill-health | No | 0 | 770 | 4 (0.5) | 774 | 0.101 |
| | Yes | | 20454 | 238 (1.2) | 20692 | |
| Had a deworming event | No | 0 | 3285 | 30 (0.9) | 3315 | 0.187 |
| | Yes | | 17939 | 212 (1.2) | 18151 | |
| Last deworming event date | 1-3 months | 3428 | 9024 | 133 (1.5) | 9157 | <0.001 |
| | >3 months | | 8802 | 79 (0.9) | 8881 | |
| Jigger treatment event | No | 12687 | 7304 | 147 (2.0) | 7451 | 0.225 |
| | Yes | | 1295 | 33 (2.5) | 1328 | |
| Hygiene promotion | No | 78 | 1841 | 27 (1.5) | 1868 | 0.179 |
| | Yes | | 19305 | 215 (1.1) | 19520 | |
| Jigger prevention program | No | 113 | 19104 | 192 (1.0) | 19296 | <0.001 |
| | Yes | | 2007 | 50 (2.4) | 2057 | |
| **Mean (sd)** | | | | | | |
| Pupil age | | 0 | 10.8 (2.0) | 10.1 (1.8) | 10.8 (2.0) | <0.001 |
| Headteacher age | | 73 | 48.7 (7.7) | 50.3 (6.7) | 48.7 (7.9) | 0.001 |
| Headteacher duration in school | | 113 | 4.2 (4.0) | 4.4 (4.9) | 4.2 (4.1) | 0.505 |
| Ratio of boys: girls | | 110 | 1.08 (0.23) | 1.13 (0.27) | 1.07 (0.23) | 0.001 |
| Ratio of pupils: teachers | | 0 | 37.4 (20.5) | 44.4 (24.6) | 37.3 (20.6) | <0.001 |
| Ratio of boys: male toilets | | 0 | 40.6 (35.3) | 40 (31.8) | 40.6 (35.3) | 0.776 |
| Ratio of girls: female toilets | | 87 | 34.8 (30.1) | 32.4 (28.9) | 34.8 (30.1) | 0.205 |

#### S2 Distribution of pupil population by classroom variables

| Variables | Categories | Number missing | Uninfected Pupils N | Infected Pupils N (%) | Total | P-value |
|---|---|---|---|---|---|---|
| All | | | 5050 | 56 (1.1) | 5106 | |
| County | Turkana | 0 | 693 | 3 (0.4) | 696 | <0.001 |
| | Samburu | | 460 | 6 (1.3) | 466 | |
| | Kericho | | 804 | 5 (0.6) | 809 | |
| | Muranga | | 665 | 20 (2.9) | 685 | |
| | Nakuru | | 879 | 6 (0.7) | 885 | |
| | Kajiado | | 632 | 4 (0.6) | 636 | |
| | Makueni | | 652 | 5 (0.8) | 657 | |
| | Taita taveta | | 112 | 0 | 112 | |
| | Kilifi | | 153 | 7 (4.4) | 160 | |
| Pupil sex | Female | | 2564 | 18 (0.7) | 2582 | 0.006 |
| | Male | | 2485 | 38 (1.5) | 2523 | |
| Roof materials | Iron sheet | 39 | 4980 | 56 (1.1) | 5036 | 0.840 |
| | Natural | | 14 | 0 | 14 | |
| | other | | 17 | 0 | 17 | |
| Wall materials | Stone/brick | 53 | 4827 | 52 (1.1) | 4879 | 0.171 |
| | other | | 174 | 0 | 174 | |
| Floor materials | Concrete | 16 | 4994 | 56 | 5050 | 0.503 |
| | other | | 40 | 0 | 40 | |
| Condition of concrete floors | Good | 230 | 2195 | 17 (0.8) | 2212 | 0.399 |
| | Slightly cracked | | 2099 | 23 (1.1) | 2122 | |
| | Very broken | | 532 | 7 (1.3) | 539 | |
| Amount of loose dust/soil | None | 4 | 1232 | 4 (0.3) | 1236 | <0.001 |
| | A little | | 3059 | 27 (0.9) | 3086 | |
| | A lot | | 755 | 25 (3.2) | 780 | |
| Classroom cleanliness | Tidy & clean | 0 | 1618 | 9 (0.6) | 1627 | <0.001 |
| | Slightly tidy & clean | | 2868 | 26 (0.9) | 2894 | |
| | Very untidy & dirty | | 564 | 21 (3.4) | 585 | |
| **Mean (sd)** | | | | | | |
| Pupil age | | | 10.8 (1.9) | 10.1 (1.6) | 10.8 (1.9) | 0.004 |
| No. pupils enrolled in class | | 35 | 37.9 (17.5) | 43.0 (21.4) | 38.0 (17.5) | 0.032 |
| No. pupils absent on the day | | 0 | 2.0 (3.2) | 3.7 (4.7) | 2.0 (3.2) | <0.001 |
| Pupil density (pupils / $m^2$) | | 17 | 0.7 (0.5) | 0.6 (0.3) | 0.7(0.5) | 0.037 |

#### S3 Correlation of classroom variables with each other

| Number of pupils | **Amount of loose dust soil** | | | |
|---|---|---|---|---|
| **Concrete floor condition** | None | A little | A lot | Total |
| Good | 959 | 1,059 | 194 | 2,212 |
| Slightly cracked | 214 | 1,725 | 183 | 2,122 |
| Very broken | 31 | 191 | 317 | 539 |
| Total | 1,204 | 2,975 | 694 | 4,873 |

**Pearson chi2(4) = 1.7e+03 P<0.001**

| Number of pupils | **Classroom cleanliness** | | | |
|---|---|---|---|---|
| **Concrete floor condition** | Tidy & clean | Slightly tidy & clean | Very untidy & dirty | Total |
| Good | 1,190 | 918 | 104 | 2,212 |
| Slightly cracked | 325 | 1,586 | 211 | 2,122 |
| Very broken | 28 | 310 | 201 | 539 |
| Total | 1,543 | 2,814 | 516 | 4,873 |

**Pearson chi2(4) = 1.3e+03 P< 0.001**

| Number of pupils | **Amount of loose dust soil** | | | |
|---|---|---|---|---|
| **Classroom cleanliness** | none | A little | A lot | Total |
| Tidy & clean | 842 | 785 | 0 | 1,627 |
| Slightly tidy & clean | 374 | 2,150 | 366 | 2,890 |
| Very untidy & dirty | 20 | 151 | 414 | 585 |
| Total | 1,236 | 3,086 | 780 | 5,102 |

**Pearson chi2(4) = 2.5e+03 P< 0.001**
